# Design of Learner-Created Electronic Flashcards in Undergraduate Medical Education: Implications for Knowledge and Skill Development

**DOI:** 10.1101/2025.09.11.25335573

**Authors:** Asavari Rajpurkar, Philip D Barrison, Kirsten Fiestan, Pratik S Vadlamudi, Matthew Sparling, Zach Landis-Lewis, Alexandra H Vinson, Emily A Balczewski

## Abstract

EFs are an increasingly popular active learning tool among medical learners. Many learners use EFs created by other learners—the content and quality of which may go unchecked by experts and educators. Of central concern is whether learner-created flashcards adequately support competency and skill development in addition to facilitating knowledge acquisition. Our mixed-methods study applied an inductively and deductively developed framework for EF design to a random sample of 1,179 flashcards from popular learner-created EF decks for undergraduate medical education (UME). We categorized each flashcard across four different dimensions: Content Area (e.g., flashcard is relevant to clinical vs. basic science fields), Clinical Reasoning Domain (e.g., flashcard supports reasoning like diagnosis), Knowledge Domain (e.g., flashcard tests procedural vs. informational knowledge), and Cognitive Process (e.g., flashcard involves recalling vs. applying knowledge). EFs in our sample tested a wide variety of knowledge in a variety of ways beyond simple factual recall. We report learner-created EFs can likely elicit higher-order thinking skills and test clinically relevant knowledge which may support clinical reasoning and procedural skill development. However, while Step 2 decks contained more clinically relevant and complex flashcards than Step 1 or preclinical decks, overall clinical relevance and complexity in learner-created EFs was limited. EFs have the potential to support learners in both knowledge acquisition and development of critical skills and competencies. However, learner-created EF decks may benefit from oversight by expert-educators to ensure their content and quality aligns with educational goals.

## Introduction

In recent years, medical learners have adapted their learning to include a variety of third-party, “parallel curriculum” study resources [1–3]. Because these resources exist outside of school-sanctioned curricula, many educators may be “unaware of the types and amounts… [that] students are using” [4]. One such parallel curriculum resource, electronic flashcards (EFs), have emerged as a highly popular active learning tool, with some studies showing adoption rates in undergraduate medical education (UME) cohorts as high as 85%, rivaling the use of traditional curricular resources like lectures [5,6]. However, unlike curricular resources, EFs are often developed and maintained by learners themselves [7] For example, several studies suggest that learners typically use EF decks which have been crowdsourced by other learners and distributed digitally on software platforms like Anki [4,8–11] As such, popular EF decks may not have been “vetted for either quality or content by the medical school establishment” [7,12]. To meet Liaison Committee on Medical Education (LCME) requirements for educational oversight and robustly support their learners, medical educators must explore what knowledge is tested and how that knowledge is tested in learner-created EFs [7].

Clarifying the educational quality and content of learner-created EFs is critical to define their role alongside other learning modalities in medical education. In particular, EFs should be evaluated for their potential to close the classroom-clinic gap, wherein clinical aptitude—not exam scores or other measures of academic performance—is the primary benchmark of success [13]. For a resource to support clinical performance, the resource should engage the types of knowledge and ways of thinking routinely required of physicians [14]. For example, resources should prepare students for clinical reasoning tasks such as diagnosis, which requires knowledge about diseases and their presentations as well as cognitive as well as procedural skills to gather and act upon information in the clinical context [14]. EFs, including learner-created EFs, have largely been shown in literature as effective in facilitating the former: knowledge acquisition as reflected by exam scores [15]. However, can EFs contribute to development of complex cognitive and procedural skills which underlie clinical performance?

Several studies suggest that expert-created EF decks may engage with such skills. Spicer et al created curriculum-aligned EFs that “promoted higher order cognitive processes, like explaining concepts, applying content to new problems, and comparing/contrasting course material” [16]. Such higher-order thinking skills (HOTS) have also been incorporated into EF creation by other groups of experts/educators [17–20]. Additionally, Schmidmeier et al found that an expert-created EF deck could facilitate acquisition and application of procedural knowledge in addition to factual or informational knowledge [21]. However, no studies, to our understanding, examine whether commonly used, learner-created decks might contain comparable or appropriate complexity. In fact, Spicer et al pursued expert curation of EFs because they noted that EFs created by learners at their US medical school too often focused on “factual recall” and “unimportant minutiae” [16].

EFs are an increasingly popular parallel curriculum resource among medical learners. Existing research on EFs has largely examined whether—not how—EFs work. In this study, we explore whether popular, learner-created EFs might promote skill development in addition to knowledge acquisition. Using a theory-informed, inductively and deductively developed framework [22], this mixed-methods study quantifies the types of content, knowledge, and cognitive skills which may be facilitated by EFs and their relevance to clinical reasoning tasks.

## Method

Our study was determined exempt from ongoing review by the University of Michigan IRBMED (HUM00222432).

### Sampling

Our study sampled six Anki decks chosen for their routine use in UME. Five decks (all but the UMMS Preclinical deck) are top recommendations in the popular “Med School Anki” community on the social media website Reddit [23], and two of these (AnKing Step 1 and AnKing Step 2) have been reported in prior studies of EF use [24,25]. The final deck (UMMS Preclinical) is commonly used at our institution. The study decks focus on three different phases of UME: preclinical curriculum (UMMS Preclinical and Foundations), Step 1 exam (AnKing Step 1 and Dope Medical Science), and Step 2 exam (AnKing Step 2 and Dope Clinical Science). Further details about the structure, size, and origin of these decks may be found in Table 1. Within each deck, we randomly sampled 150 notes, where “note” is the terminology in the Anki platform for the base unit of flashcard creation. Because a single note can give rise to multiple flashcards, the number of flashcards analyzed in each deck ranged between 164 and 264. Three flashcards in the Foundations deck have been excluded from subsequent analysis, because they simply provided information and did not ask a question, and, therefore, many aspects of our framework did not apply.

**Table 1.** Study Deck Information. Table originally published in prior manuscript [22], reproduced here for ease of reference.

| Deck Name | Component Subdecks | Curricular Phase | Country of Origin | Crowd-sourced | Number of Cards | Download Source | Download Date | Upload Date | Deck Version |
| --- | --- | --- | --- | --- | --- | --- | --- | --- | --- |
| AnKing Step 1 | Lolnotacop, Zanki Step 1 & Pharmacology | Step 1 | USA | X | 31475 | Reddit -> MEGA Drive | 11 January 2023 | 19 Feb 2022 | 11 |
| AnKing Step 2 | Zanki Step 2, Cheesy Dorian, MedicalArk | Step 2 | USA | X | 11777 | Reddit -> MEGA Drive | 11 January 2023 | 19 Feb 2022 | 11 |
| Dope: Medical Science | Bros | Step 1 | Australia |  | 21933 | Reddit -> Google Drive | 26 October 2022 | 1 May 2018 | 1 |
| Dope: Clinical Medicine | Bros | Step 2 | Australia |  | 15752 | Reddit -> Google Drive | 26 October 2022 | 1 May 2018 | 1 |
| Foundations |  | Preclinical - 2 year | USA |  | 15154 | Reddit -> Google Drive | 26 October 2022 | 21 Jul 2019 | 1 |
| UMMS Preclinical |  | Preclinical - 1 year | USA |  | 28165 | N/A | 16 January 2023 | 2020-2021 | 1 |

### EF Characteristics

This study quantified 4 different categories of EF characteristics: Content Area, Clinical Reasoning Domain, Knowledge Domain, and Cognitive Process.

The development of the latter two dimensions of EF characteristics—Knowledge Domain and Cognitive Process—through application of the *New Taxonomy of Educational Objectives* (*NT*) by Marzano and Kendall [26] is comprehensively discussed in our prior qualitative research study [22]. The aim of this prior study was to identify the design features in electronic flashcards for medical education. The study leveraged the same 6 decks used in this study as substrate for a flexible approach to thematic analysis and resulted in the Design Elements of Anki for Medical Education (DEAME) Framework which outlines seven categories that describe overarching themes of flashcard design [22].

The remaining two categories of EF characteristics quantified in this study—Content Area and Clinical Reasoning Domain—were analogously developed during our prior qualitative research study, but were excluded from the final DEAME Framework because they are features of knowledge, not elements of EF design. Therefore, specifics regarding the development of Content Area and Clinical Reasoning Domain beyond what is described in the prior study are described in Results below.

### Framework Quantification

Once thematic saturation was determined to have been reached in the prior qualitative research study to develop the DEAME Framework, a quantification of framework categories was undertaken by EB and AR on a novel sample of EFs [27]. Notably, EB and AR participated critically throughout the prior qualitative study and had substantial experience with framework categories and their application to EFs. EB and AR each coded half of the study sample independently and then met to reconcile code applications. Through discussion, they were able to reach consensus on all code applications without necessitating substantial changes to framework definitions. Codes were applied in the Dedoose qualitative coding software and analyzed in the R programming language [28,29]. For each EF deck, we report the number and percentage of flashcards with a given characteristic. When comparing groups of decks (e.g., preclinical vs. Step 1 decks) during analysis, we report the mean of percentages for each group.

## Results

A total of 1,179 electronic flashcards were analyzed across six commonly used EF decks for UME. The quantification of 4 different categories of EF characteristics—Content Area, Clinical Reasoning Domain, Knowledge Domain, and Cognitive Process—can be found in Table 2. Results for each category and specifics about category development are discussed in the following sections.

**Table 2.** Quantification of EF Characteristics.

|  | <u>Preclinical</u> |  | <u>Step 1</u> |  | <u>Step 2</u> |  |
| --- | --- | --- | --- | --- | --- | --- |
| **Characteristic** | **UMMS**<br>**Preclinical**<br>**N = 211** | **Foundations**<br>**N = 264\*** | **AnKing**<br>**Step 1**<br>**N = 189** | **Dope Medical**<br>**Science**<br>**N = 164** | **AnKing**<br>**Step 2**<br>**N = 170** | **Dope**<br>**Clinical**<br>**Science**<br>**N = 181** |
| ***Primary Content Area<sup>a</sup>*** |  |  |  |  |  |  |
| Basic Science Field | 152 (72%) | 237 (90%) | 99 (52%) | 94 (57%) | 40 (24%) | 55 (30%) |
| Clinical Field | 59 (28%) | 27 (10%) | 90 (48%) | 70 (43%) | 130 (76%) | 126 (70%) |
| ***Clinical Reasoning Domain<sup>b</sup>*** |  |  |  |  |  |  |
| Diagnosis | 37 (18%) | 24 (9%) | 35 (19%) | 37 (23%) | 66 (39%) | 63 (35%) |
| Etiology or Risk Factors | 18 (9%) | 6 (2%) | 35 (19%) | 36 (22%) | 17 (10%) | 34 (19%) |
| Management | 13 (6%) | 3 (1%) | 19 (10%) | 5 (3%) | 47 (28%) | 29 (16%) |
| Prognosis | 11 (5%) | 4 (1%) | 17 (9%) | 12 (7%) | 21 (12%) | 9 (5%) |
| Ethics | 0 (0%) | 1 (0%) | 0 (0%) | 0 (0%) | 0 (0%) | 0 (0%) |
| None | 132 (63%) | 226 (86%) | 83 (44%) | 74 (45%) | 19 (11%) | 46 (25%) |
| ***Knowledge Domain<sup>c</sup>*** |  |  |  |  |  |  |
| Information | 185 (88%) | 255 (97%) | 170 (90%) | 157 (96%) | 118 (69%) | 131 (72%) |
| Mental Procedure | 24 (11%) | 8 (3%) | 19 (10%) | 7 (4%) | 52 (31%) | 47 (26%) |
| Psychomotor Procedure | 2 (1%) | 1 (0%) | 0 (0%) | 0 (0%) | 0 (0%) | 3 (2%) |
| ***Cognitive Process<sup>d</sup>*** |  |  |  |  |  |  |
| Retrieval | 161 (76%) | 241 (91%) | 165 (87%) | 135 (82%) | 122 (72%) | 130 (72%) |
| Comprehension | 9 (4%) | 4 (2%) | 0 (0%) | 7 (4%) | 1 (1%) | 1 (1%) |
| Analysis | 40 (19%) | 16 (6%) | 23 (12%) | 21 (13%) | 26 (15%) | 36 (20%) |
| Knowledge Utilization | 1 (0%) | 3 (1%) | 1 (1%) | 1 (1%) | 21 (12%) | 14 (8%) |

Cells colored according to percentage by deck within each of the four categories of EF characteristics.
<sup>a</sup>Primary Content Area = the EF was most pertinent to a field with Basic Science (16 total fields, e.g., pathology) vs. Clinical (29 total fields, e.g., neurology) focus
<sup>b</sup>Clinical Reasoning Skill = the EF contained information directly supportive for executing a clinical reasoning task
<sup>c</sup>Knowledge Domain = the EF tested knowledge in any of three categories derived from the *New Taxonomy of Educational Objectives* by Marzano and Kendall [26]
<sup>d</sup>Cognitive Process = the EF likely elicited thinking process in any of four categories derived from the *New Taxonomy of Educational Objectives* by Marzano and Kendall [26]

### Content Area

Given that EF decks in our study targeted complete preclinical curricula or summative exams, EFs in our study included a variety of content areas. Content areas were identified iteratively and inductively when flashcards with novel content were encountered. A total of 45 content areas were identified, named according to standard terminology (e.g., medical fellowships) where appropriate, and grouped into Basic Science or Clinical Fields. Fields which are established areas of clinical practice were classified as Clinical Fields (n = 29), excepting Histology and Pathology, Nutrition, Radiology, and Toxicology which were included as Basic Science Fields because they are cross-cutting, foundational, and often incorporated early in preclinical curricula. The remaining fields were classified as Basic Science Fields (n = 16).

As seen in Table 3, the most common Basic Science Fields among all decks were Anatomy (mean = 17%), Histology and Pathology (mean = 14%), and Pharmacology (mean = 10%) and the most common Clinical Fields across all decks were Neurology (mean = 16%), Gastroenterology (mean = 13%), and Infectious Disease (mean = 12%). The fields of Internal Medicine, Family Medicine, and Geriatrics were collapsed into a single code of exclusion called “General Adult Medicine” due to their broad applicability and substantial overlap with one another. As such, relevant subspecialties like Cardiology may be combined with General Adult Medicine to roughly correlate the relevance of our study sample to these general fields. For example, 83% of all flashcards in our study sample may pertain to Internal Medicine (IM) after combining the observed frequencies of General Adult Medicine and 11 common IM subspecialties (Allergy and Immunology, Cardiology, Clinical Genetics, Endocrinology, Gastroenterology, Hematology, Infectious Disease, Nephrology, Oncology, Pulmonology and Acute Care, Rheumatology; Table 1 in Online Resource 1).

**Table 3.** Content Areas in Study Decks.

|  | <u>Preclinical</u> |  |  |  | <u>Step 1</u> |  |  |  | <u>Step 2</u> |  |  |  |
| --- | --- | --- | --- | --- | --- | --- | --- | --- | --- | --- | --- | --- |
| Content Area | UMMS<br>Preclinical<br>N = 211 |  | Foundations<br>N = 264* |  | AnKing<br>Step 1<br>N = 189 |  | Dope Medical Science<br>N = 164 |  | AnKing<br>Step 2<br>N = 170 |  | Dope Clinical Science<br>N = 181 |  |
| *Basic Science Field* | *Any* | *Primary* | *Any* | *Primary* | *Any* | *Primary* | *Any* | *Primary* | *Any* | *Primary* | *Any* | *Primary* |
| Anatomy | 27 (13%) | 21 (11%) | 150 (57%) | 129 (51%) | 16 (8%) | 10 (5%) | 19 (12%) | 12 (8%) | 4 (2%) | 0 (0%) | 17 (9%) | 15 (9%) |
| Biochemistry | 28 (13%) | 12 (7%) | 4 (2%) | 4 (2%) | 15 (8%) | 8 (4%) | 14 (9%) | 7 (5%) | 3 (2%) | 3 (2%) | 3 (2%) | 0 (0%) |
| Biostatistics | 2 (1%) | 2 (1%) | 5 (2%) | 5 (2%) | 1 (1%) | 1 (1%) | 0 (0%) | 0 (0%) | 0 (0%) | 0 (0%) | 0 (0%) | 0 (0%) |
| Cell Biology | 13 (6%) | 0 (0%) | 10 (4%) | 0 (0%) | 15 (8%) | 8 (4%) | 8 (5%) | 6 (4%) | 0 (0%) | 0 (0%) | 1 (1%) | 1 (1%) |
| Embryology | 5 (2%) | 5 (3%) | 0 (0%) | 0 (0%) | 3 (2%) | 1 (1%) | 4 (2%) | 4 (3%) | 2 (1%) | 0 (0%) | 0 (0%) | 0 (0%) |
| Genetics | 7 (3%) | 3 (2%) | 2 (1%) | 0 (0%) | 8 (4%) | 1 (1%) | 9 (5%) | 4 (3%) | 0 (0%) | 0 (0%) | 2 (1%) | 0 (0%) |
| Health Systems and Safety | 0 (0%) | 0 (0%) | 13 (5%) | 11 (4%) | 0 (0%) | 0 (0%) | 0 (0%) | 0 (0%) | 0 (0%) | 0 (0%) | 0 (0%) | 0 (0%) |
| Histology and Pathology | 37 (18%) | 27 (15%) | 49 (19%) | 35 (14%) | 26 (14%) | 12 (6%) | 28 (17%) | 10 (7%) | 14 (8%) | 1 (1%) | 13 (7%) | 7 (4%) |
| Immunology | 17 (8%) | 10 (5%) | 4 (2%) | 0 (0%) | 4 (2%) | 3 (2%) | 8 (5%) | 2 (1%) | 2 (1%) | 0 (0%) | 2 (1%) | 1 (1%) |
| Microbiology | 14 (7%) | 5 (3%) | 3 (1%) | 2 (1%) | 21 (11%) | 17 (9%) | 16 (10%) | 9 (6%) | 4 (2%) | 2 (1%) | 1 (1%) | 0 (0%) |
| Nutrition | 1 (0%) | 0 (0%) | 0 (0%) | 0 (0%) | 6 (3%) | 4 (2%) | 2 (1%) | 1 (1%) | 1 (1%) | 1 (1%) | 1 (1%) | 0 (0%) |
| Pharmacology | 23 (11%) | 20 (11%) | 3 (1%) | 1 (0%) | 26 (14%) | 18 (10%) | 22 (13%) | 19 (12%) | 21 (12%) | 8 (5%) | 12 (7%) | 4 (3%) |
| Physiology | 33 (16%) | 19 (10%) | 18 (7%) | 15 (6%) | 19 (10%) | 11 (6%) | 13 (8%) | 7 (5%) | 2 (1%) | 1 (1%) | 8 (4%) | 3 (2%) |
| Public Health | 1 (0%) | 1 (1%) | 9 (3%) | 5 (2%) | 11 (6%) | 1 (1%) | 6 (4%) | 1 (1%) | 4 (2%) | 0 (0%) | 5 (3%) | 2 (1%) |
| Radiology | 0 (0%) | 0 (0%) | 24 (9%) | 17 (7%) | 1 (1%) | 1 (1%) | 5 (3%) | 1 (1%) | 12 (7%) | 6 (4%) | 5 (3%) | 1 (1%) |
| Toxicology | 0 (0%) | 0 (0%) | 2 (1%) | 2 (1%) | 2 (1%) | 0 (0%) | 1 (1%) | 0 (0%) | 0 (0%) | 0 (0%) | 0 (0%) | 0 (0%) |
| *Clinical Field* | *Any* | *Primary* | *Any* | *Primary* | *Any* | *Primary* | *Any* | *Primary* | *Any* | *Primary* | *Any* | *Primary* |
| Allergy and Immunology | 1 (0%) | 0 (0%) | 0 (0%) | 0 (0%) | 2 (1%) | 0 (0%) | 0 (0%) | 0 (0%) | 1 (1%) | 0 (0%) | 0 (0%) | 0 (0%) |
| Anesthesiology | 1 (0%) | 0 (0%) | 2 (1%) | 0 (0%) | 0 (0%) | 0 (0%) | 0 (0%) | 0 (0%) | 3 (2%) | 2 (1%) | 2 (1%) | 0 (0%) |
| Cardiology | 27 (13%) | 4 (2%) | 7 (3%) | 0 (0%) | 17 (9%) | 5 (3%) | 7 (4%) | 3 (2%) | 26 (15%) | 11 (7%) | 22 (12%) | 14 (9%) |
| Clinical Genetics | 2 (1%) | 1 (1%) | 0 (0%) | 0 (0%) | 3 (2%) | 1 (1%) | 1 (1%) | 0 (0%) | 6 (4%) | 0 (0%) | 1 (1%) | 1 (1%) |
| Dentistry | 0 (0%) | 0 (0%) | 0 (0%) | 0 (0%) | 3 (2%) | 0 (0%) | 1 (1%) | 1 (1%) | 0 (0%) | 0 (0%) | 0 (0%) | 0 (0%) |
| Dermatology | 20 (9%) | 1 (1%) | 10 (4%) | 4 (2%) | 10 (5%) | 4 (2%) | 4 (2%) | 4 (3%) | 11 (6%) | 4 (3%) | 8 (4%) | 2 (1%) |
| Emergency Medicine | 0 (0%) | 0 (0%) | 0 (0%) | 0 (0%) | 0 (0%) | 0 (0%) | 0 (0%) | 0 (0%) | 1 (1%) | 0 (0%) | 1 (1%) | 0 (0%) |
| Endocrinology | 12 (6%) | 5 (3%) | 13 (5%) | 0 (0%) | 17 (9%) | 10 (5%) | 14 (9%) | 1 (1%) | 19 (11%) | 6 (4%) | 8 (4%) | 3 (2%) |
| Gastroenterology | 22 (10%) | 1 (1%) | 15 (6%) | 0 (0%) | 21 (11%) | 9 (5%) | 17 (10%) | 5 (3%) | 23 (14%) | 8 (5%) | 49 (27%) | 20 (13%) |
| General Adult Medicine | 0 (0%) | 0 (0%) | 2 (1%) | 1 (0%) | 3 (2%) | 1 (1%) | 0 (0%) | 0 (0%) | 4 (2%) | 1 (1%) | 7 (4%) | 2 (1%) |
| General and Trauma Surgery | 12 (6%) | 4 (2%) | 53 (20%) | 0 (0%) | 4 (2%) | 2 (1%) | 2 (1%) | 1 (1%) | 19 (11%) | 13 (9%) | 29 (16%) | 21 (13%) |
| Pediatrics | 2 (1%) | 1 (1%) | 5 (2%) | 0 (0%) | 10 (5%) | 1 (1%) | 5 (3%) | 1 (1%) | 16 (9%) | 6 (4%) | 16 (9%) | 9 (6%) |
| Hematology | 14 (7%) | 6 (3%) | 13 (5%) | 2 (1%) | 28 (15%) | 5 (3%) | 27 (16%) | 9 (6%) | 17 (10%) | 4 (3%) | 11 (6%) | 2 (1%) |
| Infectious Disease | 19 (9%) | 13 (7%) | 4 (2%) | 0 (0%) | 32 (17%) | 10 (5%) | 26 (16%) | 11 (7%) | 28 (16%) | 12 (8%) | 20 (11%) | 3 (2%) |
| Nephrology | 19 (9%) | 1 (1%) | 7 (3%) | 0 (0%) | 14 (7%) | 2 (1%) | 8 (5%) | 1 (1%) | 11 (6%) | 4 (3%) | 4 (2%) | 1 (1%) |
| Neurology | 27 (13%) | 4 (2%) | 59 (22%) | 5 (2%) | 31 (16%) | 6 (3%) | 23 (14%) | 9 (6%) | 18 (11%) | 8 (5%) | 38 (21%) | 8 (5%) |
| Neurosurgery | 0 (0%) | 0 (0%) | 33 (13%) | 0 (0%) | 1 (1%) | 0 (0%) | 3 (2%) | 1 (1%) | 3 (2%) | 0 (0%) | 8 (4%) | 3 (2%) |
| Obstetrics and Gynecology | 15 (7%) | 5 (3%) | 32 (12%) | 3 (1%) | 13 (7%) | 5 (3%) | 8 (5%) | 3 (2%) | 29 (17%) | 14 (9%) | 9 (5%) | 2 (1%) |
| Oncology | 7 (3%) | 3 (2%) | 10 (4%) | 7 (3%) | 21 (11%) | 11 (6%) | 33 (20%) | 15 (10%) | 16 (9%) | 15 (10%) | 19 (10%) | 8 (5%) |
| Ophthalmology | 8 (4%) | 0 (0%) | 3 (1%) | 0 (0%) | 1 (1%) | 1 (1%) | 0 (0%) | 0 (0%) | 3 (2%) | 1 (1%) | 4 (2%) | 3 (2%) |
| Orthopedic Surgery | 6 (3%) | 1 (1%) | 43 (16%) | 1 (0%) | 11 (6%) | 2 (1%) | 5 (3%) | 0 (0%) | 6 (4%) | 4 (3%) | 6 (3%) | 4 (3%) |
| Otolaryngology | 6 (3%) | 0 (0%) | 37 (14%) | 0 (0%) | 7 (4%) | 2 (1%) | 2 (1%) | 0 (0%) | 1 (1%) | 1 (1%) | 9 (5%) | 6 (4%) |
| Pediatric Surgery | 0 (0%) | 0 (0%) | 0 (0%) | 0 (0%) | 0 (0%) | 0 (0%) | 0 (0%) | 0 (0%) | 0 (0%) | 0 (0%) | 1 (1%) | 0 (0%) |
| Physical Medicine and Rehabilitation | 0 (0%) | 0 (0%) | 0 (0%) | 0 (0%) | 1 (1%) | 0 (0%) | 0 (0%) | 0 (0%) | 0 (0%) | 0 (0%) | 0 (0%) | 0 (0%) |
| Plastic Surgery | 0 (0%) | 0 (0%) | 0 (0%) | 0 (0%) | 0 (0%) | 0 (0%) | 0 (0%) | 0 (0%) | 0 (0%) | 0 (0%) | 0 (0%) | 0 (0%) |
| Psychiatry | 11 (5%) | 0 (0%) | 14 (5%) | 4 (2%) | 7 (4%) | 2 (1%) | 7 (4%) | 2 (1%) | 9 (5%) | 3 (2%) | 7 (4%) | 2 (1%) |
| Pulmonology and Acute Care | 11 (5%) | 5 (3%) | 13 (5%) | 0 (0%) | 18 (10%) | 3 (2%) | 10 (6%) | 1 (1%) | 25 (15%) | 5 (3%) | 25 (14%) | 10 (6%) |
| Rheumatology | 14 (7%) | 4 (2%) | 0 (0%) | 0 (0%) | 9 (5%) | 7 (4%) | 9 (5%) | 2 (1%) | 13 (8%) | 5 (3%) | 6 (3%) | 1 (1%) |
| Urology | 8 (4%) | 0 (0%) | 13 (5%) | 0 (0%) | 6 (3%) | 0 (0%) | 2 (1%) | 0 (0%) | 4 (2%) | 0 (0%) | 10 (6%) | 0 (0%) |

Most flashcards in our sample (86%) contained more than one relevant content area. Therefore, we additionally identified the most prominent (i.e., “primary”) content area among these. When making this determination, we considered the direct clinical applicability of the flashcard’s content. For example, a flashcard which tested the mechanism of action of a diabetes drug should have a primary content area of Pharmacology (a Basic Science Field) while a flashcard which tested the clinical context in which that drug should be prescribed (i.e., diabetes) should have a primary content area of Endocrinology (a Clinical Field). As seen in Table 2, preclinical decks largely had a Basic Science Field as a primary content area (mean Primary Basic Science Field = 81%; mean Primary Clinical Field = 19%), while Step 1 decks had a roughly even representation of primary content areas (mean Primary Basic Science Field = 55%, mean Primary Clinical Field = 46%) (Figure 1). Step 2 decks predominantly pertained to Clinical Fields (mean Primary Basic Science Field = 27%, mean Primary Clinical Field = 73%).

### Clinical Reasoning Domain

To support clinical reasoning, physicians have “information needs” relating to the following mutually exclusive categories—Diagnosis, Etiology, Management, and Prognosis— which we call Clinical Reasoning Domains [30–32]. To clarify the boundaries between these categories, we considered the category Etiology to include risk factors in addition to known cause-effect relationships, Management to include preventative management in addition to reactive management, and Prognosis to include complications resulting from an intervention in addition to outcomes resulting from a disease process. Additionally, we added a further category—Ethics—after encountering a single flashcard in the Foundations preclinical deck which engaged with ethical principles foundational to clinical reasoning. Notably, to be assigned a Clinical Reasoning Domain, a flashcard need not require the completion of a clinical reasoning task, but simply provide information directly relevant to its execution. For example, a flashcard which correlated a histological finding to a disease would support “Diagnosis,” without necessitating that the user diagnose that disease from, e.g., a clinical vignette containing that histological finding. Likewise, a flashcard which tested general steps for histological analysis without correlation to a specific disease would not be assigned a Clinical Reasoning Domain. All decks contained flashcards (mean = 54%) that furnished information supportive of clinical reasoning (Table 2). Step 2 decks had the highest percentage of flashcards relevant to Clinical Reasoning Domains (mean = 82%), followed by Step 1 (mean = 55%) and preclinical decks (mean = 25%). The most common Clinical Reasoning Domains were Diagnosis (mean = 24%) and Etiology (mean = 14%).

### Knowledge Domain

The Knowledge Domain category is adapted from the *NT* and describes three types of knowledge which a flashcard might involve: Information, Mental Procedural, and Psychomotor Procedural knowledge [22,26]. Information describes “characteristics” of and “relationships” between “persons, places, living and nonliving things, and events” [26]. Most flashcards across all decks (mean = 85%) tested Informational knowledge. Therefore, a minority of flashcards across all decks (mean = 15%) tested Procedural knowledge which describes how or why a task or action should be performed [26]. Notably, however, almost 1/3 of flashcards in the Step 2 decks (mean = 30%) incorporated Procedural knowledge, in contrast to preclinical (mean = 8%) and Step 1 (mean = 7%) decks. Across all decks, Mental Procedures (mean = 14%), which include cognitive actions like diagnosis and management, were remarkably more common than Psychomotor Procedures (mean = 1%), which include physical actions like clinical exam maneuvers.

### Cognitive Process

The Cognitive Process category is also adapted from the NT and describes the type of thinking a learner might engage to generate an EF answer [26]. The four codes are roughly ordered by cognitive complexity from the least complex, Retrieval, to the most complex, Knowledge Utilization. Retrieval requires the learner to generate an answer but not demonstrate an “in-depth” understanding of the knowledge’s “basic structure…or critical and noncritical elements” and can be considered a lower-order thinking skill (LOTS) [26]. The remaining three levels—Comprehension, Analysis, and Knowledge Utilization—can likewise be considered higher-order thinking skills (HOTS). Comprehension requires the learner to demonstrate an “in-depth” understanding of pieces of knowledge by understanding their whys or hows [26]. Finally, Analysis requires the generation of “new conclusions” from existing knowledge and Knowledge Utilization requires application of existing knowledge to “specific situations” [26]. Of note, we coded the highest Cognitive Process a flashcard might elicit when it is first seen, though multiple may be possible over time and across learners. As seen in Table 2, Retrieval (i.e., LOTS) was the most common Cognitive Process across all study decks (mean = 80%), but all decks contained a minority of HOTS (mean = 20%). HOTS were most prominent in the Step 2 decks (mean = 29%) compared to the Step 1 (mean = 16%) and preclinical decks (mean = 17%).

## Discussion

Preliminary research suggests EFs are an effective learning strategy, but little is known about the content and quality of EF decks for medical education. Of particular concern is whether popular learner-created EFs support clinical skill development, in addition to knowledge acquisition. This mixed-methods study explores the content areas, types of knowledge, and thinking skills tested in six popular learner-created Anki decks for UME. We show that EFs in our sample display significant variety across these dimensions, testing three different types of knowledge with four levels of likely elicited cognitive processes relevant to 45 clinical and basic science fields and five clinical reasoning tasks. In other words, learner-created EFs may support more than simple “factual recall” of limited content areas [16].

We find that the types of knowledge grow in clinical relevance and the ways of testing knowledge grow increasingly complex when learner-created EFs target later (i.e., the Step 2 exam) vs. earlier (i.e., the Step 1 exam and preclinical curricula) phases of the UME curriculum. In this way, learner-created EFs appear to mimic the changing goals and expectations of learners as they progress through training. However, are these learner-created EFs sufficient to support learners in achieving mastery in the clinic as well as the classroom?

Indeed, while we show that learner-created EFs in our sample *can* engage with complex and clinical content, we still find that many EFs do not. For example, over half of all EFs in our sample focused primarily on testing basic science rather than clinical content. Additionally, about half of all flashcards did not have direct relevance to clinical reasoning tasks like diagnosis and management. Similarly, while we show that EFs are able to test procedural knowledge, which can facilitate these same clinical reasoning tasks, only the Step 2 decks do this with substantial frequency. Lastly, we find that less than a quarter of EFs in our sample likely elicit HOTS like applying knowledge to make decisions in novel situations.

Thus, our data highlights an overarching challenge in medical education. Despite access to a vast amount of information, learners often remain underprepared for clinical practice and complex medical decision making because study tools prioritize rote memorization and exam performance over critical thinking and application [33]. However, previous research on problem-based learning and clinical simulation has shown that cultivating HOTS improves learners’ abilities to translate conceptual knowledge into reasoning and procedural skills [34,35]. Additionally, a large meta-analysis across STEM education has demonstrated that active learning strategies broadly reduced failure rates and improved academic performance compared with traditional lecture-based instruction, emphasizing the widespread benefits of promoting higher-level thinking skills [36].These findings underscore the value of designing study tools that intentionally promote complex reasoning skills rather than simple factual recall. Therefore, if re-structured to target HOTS, EFs could complement these established educational methods in supporting both knowledge acquisition and clinical skill development.

Several factors may explain the limited engagement of HOTS in EFs, including student preference to use EFs for rehearsing simple and factual knowledge and other learning tools for engaging with more clinical and complex concepts. However, as both learners and educators, we feel the potential of EFs to facilitate active learning of the full spectrum of knowledge and skills required of physicians should be explored. This exploration may benefit from learner-educator co-creation schemes which merge students’ enthusiasm for and knowledge of the EF format with educators’ expertise in both content and pedagogy [16]. In this way, we could better align EFs’ ability to test a variety of content in a variety of ways with the specific goals and objectives of medical education.

To enable this alignment, further work is needed to understand how EF design impacts how learners experience and learn from EFs. Indeed, a limitation of our study is that it used qualitative consensus building to map EFs to the four categories of EF used in this study but did not validate these mappings with more advanced methodologies like think-aloud studies. Were such mappings validated, templates and guidelines for flashcard design to target each of the areas in our study could be developed. Such efforts may benefit from exploration of a wider sample of EFs, including expert-created EFs or EFs on other platforms than Anki, than the limited sample our study included.

In recent years, learners have been the leaders in advancing EFs as an effective active learning tool for medical education. However, to enhance the capacity of EFs to more holistically develop our future physician workforce, we suggest that educators must partner with students in these efforts. We hope this mixed-methods study sheds light on learner-created EFs and motivates educators towards this necessary engagement with EFs.

## Data Availability

All data produced in the present study are available upon reasonable request to the authors.

## Contributions

**Asavari Rajpurkar** was responsible for the conception and design of the work, the curation of data, analysis of data, investigation of data, authorship of the initial manuscript, and revision of manuscript; and gives final approval of the version to be published.

**Philip D Barrison** was responsible for the conception and design of the work, the curation of data, analysis of data, acquisition of funding, investigation of data, design of methodology, administration of project, management of software, supervision of project, visualization of data, and revision of manuscript; and gives final approval of the version to be published.

**Kirsten Fiestan** was responsible for the analysis of data, investigation of data, and revision of manuscript; and gives final approval of the version to be published.

**Pratik S Vadlamudi** was responsible for the analysis of data, investigation of data, and revision of manuscript; and gives final approval of the version to be published.

**Matthew Sparling** was responsible for the analysis of data, investigation of data, and revision of manuscript; and gives final approval of the version to be published.

**Zach Landis-Lewis** was responsible for the conception and design of the work, acquisition of funding, design of methodology, provision of resources, supervision of project, and revision of manuscript; and gives final approval of the version to be published.

**Alexandra H Vinson** was responsible for the conception and design of the work, analysis of data, acquisition of funding, investigation of data, design of methodology, provision of resources, administration of project, supervision of project, and revision of manuscript; and gives final approval of the version to be published.

**Emily A Balczewski** was responsible for the conception and design of the work, the curation of data, analysis of data, acquisition of funding, investigation of data, design of methodology, administration of project, management of software, supervision of project, visualization of data, authorship of the initial manuscript, and revision of manuscript; and had full access to all the data in the study and takes responsibility for the integrity of the data and the accuracy of the data analysis.

## Acknowledgements

The authors wholeheartedly wish to thank Idris Nagar for uploading and inputting the metadata for our note PDFs.

## Statements and Declarations

### Informed Consent

Not applicable.

